# Protocol for the development of Pituitary Surgery Core Outcomes and Priorities (PitCOP)

**DOI:** 10.1101/2024.06.16.24308989

**Authors:** Nicola Newall, Alexandra Valetopoulou, Danyal Z Khan, Anouk Borg, Fion Bremner, Neil Dorward, Maria Fleseriu, Joy Ginn, Mark Gurnell, Marta Korbonits, Inma Serrano, Stephanie E Baldeweg, Angelos G Kolias, Hani J Marcus

**Affiliations:** Department of Neurosurgery, National Hospital for Neurology and Neurosurgery, London, United Kingdom; Wellcome/EPSRC Centre for Interventional and Surgical Sciences (WEISS), London, United Kingdom; Department of Ophthalmology, National Hospital for Neurology and Neurosurgery, London, United Kingdom; Department of Neurosurgery, Oregon Health & Science University, Portland, USA; The Pituitary Foundation, United Kingdom; Division of Neurosurgery, Department of Clinical Neurosciences, Addenbrooke’s Hospital and University of Cambridge, Cambridge, UK; Centre for Endocrinology, Barts and The London School of Medicine, Queen Mary University of London, London, United Kingdom; Department of Diabetes & Endocrinology, University College London Hospitals NHS Foundation Trust, London, United Kingdom

## Abstract

**Objectives:** We aim to (i) identify the most important research priorities in pituitary surgery through a priority setting partnership (PSP) and (ii) develop a core outcome set (COS) for pituitary surgery research.

**Design:** International modified Delphi consensus processes.

**Subjects:** Participants are key stakeholders in pituitary surgery, including: healthcare professionals (HCPs) (nurse specialists, neurosurgeons, endocrinologists, ophthalmologists, otolaryngologists, radiologists, pathologists, and oncologists); service users (patients, family members and carers); and charity representatives.

**Methods:** In PSP round one, participants are asked what questions relating to diagnosis, treatment, long-term care and follow-up they would like answered by future research. Responses will be grouped thematically, and a systematic literature review will identify which research priorities remain unanswered. In round two, participants will rank their top 10 unanswered research priorities.

In COS round one, participants are presented with an initial list of outcomes identified from relevant literature,^1,2^ and asked to (i) rate outcomes based on perceived importance for inclusion in a core outcome set, and (ii) suggest additional outcomes. In round two, participants will re-score each outcome, while considering the summarised group scores from round one.

The final round of PSP and COS will be held as a live consensus workshop (with equal representation of stakeholder groups) to determine the top 10 research priorities and the final core outcome set.

**Results:** The study is currently underway, and aims to be complete by August 2024.

**Conclusions:** The PitCOP study will establish patient-centred research priorities and a core outcome set for pituitary surgery research. This will help to improve the relevance, efficiency, and quality of future pituitary surgery research.

## Introduction

Pituitary adenomas are common benign tumours, accounting for 10 - 15% of all intracranial neoplasms^1,2^. The typical presentation of pituitary adenomas includes visual deterioration, headaches, and manifestations of hormonal imbalances. Management is guided by several factors including clinical features, endocrine profile and imaging findings, and typically involves a multidisciplinary approach. Surgical resection, via an endonasal transsphenoidal approach, is the mainstay of treatment for non-functioning pituitary adenomas (NFPA) and certain functioning adenomas.

Over recent years, there have been significant advances in pituitary adenoma surgical techniques and technology. However, despite these advances, there remain issues with complications, such as CSF rhinorrhoea and dysnatraemia, as well as post-operative outcomes, such as incomplete resection^3^. Additionally, even in patients who achieve long-term remission, it has been reported that some physical and psychological symptoms remain, adversely affecting quality of life (QoL)^4^. This impact on QoL persists even in patients who are considered to have ‘good’ outcomes by conventional metrics.

To address these challenges, research firstly needs to be focused on agreed priority areas. Current clinical research largely reflects the interests of researchers in academia or industry, which may not address the real-world needs and priorities of patients. This has therefore led to a mismatch in research priorities for patients, clinicians and researchers and the production of inefficient and ineffective research. Secondly, outcomes measured and reported should be standardized to ensure consistency and comparability across studies and enhance the reliability and applicability of findings.

One approach to align research with patients’ needs is through the use of a Priority setting partnership (PSP). A PSP aims to develop research priorities for areas of health care where there are considerable research uncertainties. A research uncertainty can be defined as any important question about a specific area of health care that cannot be convincingly answered by the existing body of evidence^5^. Research priority studies enable patients, carers, and healthcare professionals to work together to identify and prioritise the research uncertainties which are most important to them. To date, there have been a number of research priority studies covering a variety of disease areas including degenerative cervical myelopathy, breast cancer surgery and spinal cord injury^6–8^.

The use of a Core Outcome Set (COS) enables the standardization of measured outcomes. A COS is an ‘agreed standardised collection of outcomes which should be measured and reported, as a minimum, in all trials for a specific clinical area’. There are several benefits of Core Outcome Sets^9,10^:

- Reduced heterogeneity, facilitating quantitative evidence synthesis.
- Reduced selective reporting of outcomes.
- Identifying clinically relevant outcomes by involving a wide range of stakeholders, such as patients, carers, family members, multidisciplinary healthcare professionals and charity representatives.

COS have been successfully developed and implemented in various other diseases, such as rheumatoid arthritis, traumatic brain injury and spinal cord injury^11–13^.

PitCOP, therefore, aims to improve the quality, efficiency and effectiveness of future pituitary surgery research. We aim to do this by (1) identifying the most important research uncertainties in pituitary adenoma surgery from the joint perspective of patients, caregivers, and clinicians to guide future research studies and (2) developing a patient-centred core outcome set for pituitary surgery research through an international multi-stakeholder partnership.

## Methods

The PitCOP study aims to bring together stakeholders with lived or professional experience in pituitary adenoma surgery to establish a PSP and COS for use in future research studies. The key objectives of the study are: (i) to establish the top 10 research priorities (PSP); (ii) to determine which outcomes are applicable and relevant for use in future studies in pituitary adenoma surgery (COS).

### Steering Committee

A steering committee will be formed to oversee study conduct and guide development of the PSP and COS. The committee will include representatives from each stakeholder group, consisting of pituitary surgeons, endocrinologists, ophthalmologists, nurse specialists, charity representatives and service users. Steering group members will be recruited from UK and International professional and charitable organisations which include the Pituitary Society, Pituitary Foundation, Endocrine Society and European Society of Ophthalmologists. The day-to-day running of the study will be overseen by a management committee. Both groups will ensure representation from those with lived and professional experience of pituitary adenomas requiring surgery. Steering and management committee members are listed in appendix 1.

### Stakeholders

Relevant stakeholder groups include: healthcare professionals (HCPs) involved in the care of patients with pituitary adenomas (including pituitary surgeons, endocrinologists, ophthalmologists, otorhinolaryngologists, oncologists and nurse specialists); service users (including patients, their families and carers); and charity representatives from the Pituitary Foundation.

### Participant recruitment

A dedicated study webpage has been created (https://www.pit-cop.com) which contains information resources, participant registration forms and links to the surveys. The Wellcome / EPSRC Centre for Interventional and Surgical Sciences (WEISS) public engagement team have contributed to the development of participant resources. On registration respondents will provide demographics, including age, biological sex, geographic location, and stakeholder group. The first page of each online Delphi survey will be a consent form. By signing the consent form, respondents are consenting to participate, though they have the right to withdraw from the study at any stage.

Invitation to participate will be disseminated to relevant stakeholder groups by email and social media platforms. Study information will also be distributed through relevant societies including: the Pituitary Foundation (an international charity for patients with disorders of the pituitary gland), the Endocrine Society, the Pituitary Society, and the European Society of Ophthalmologists. Promotional material will be used as part of the recruitment process, including a video (appendix 2). To minimise attrition bias between Delphi rounds regular reminder emails will be sent to participants and updates regarding study progress will be posted on our social media platforms.

### Ethics and Dissemination

Ethics approval has been granted by the University of Cambridge Psychology Research Ethics Committee. The COS has been prospectively registered on the COMET database^14^.The results of this study will be published in a peer review journal and presented at conferences. Findings will also be disseminated through the PitCOP website and social media page as well as websites and social media of the relevant societies and charity organisations.

#### PSP study design

Study design and report generation were guided by the REPRISE guidelines^15^. A study timeline can be found in Figure 1.

**Figure 1:**
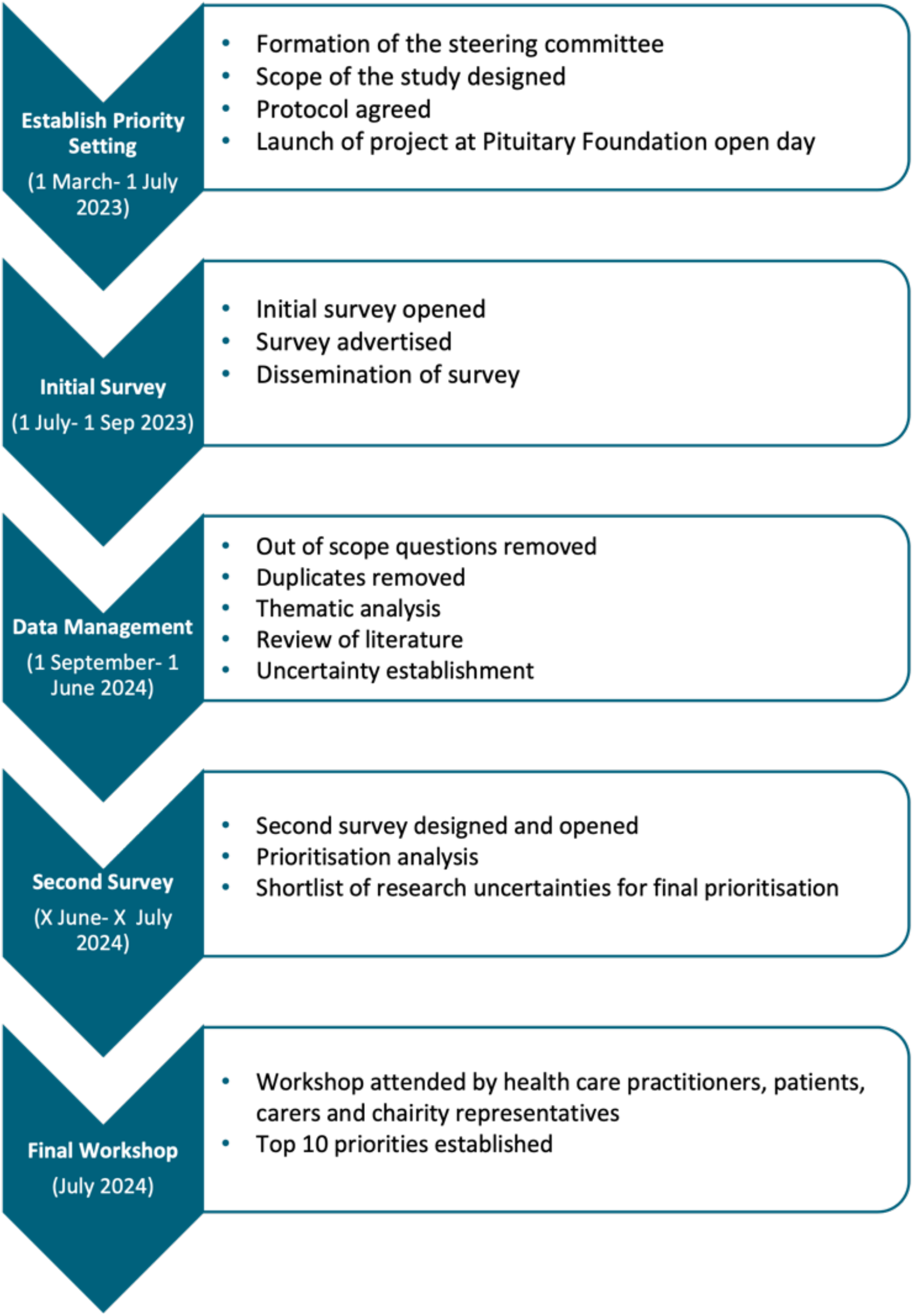
PSP study timeline

### Round 1- online survey

A multi-round online Delphi survey will be conducted on the online survey tool-*Qualtrics*. The aim of the initial survey is to gather potential research uncertainties. The survey will ask the following four questions to structure stakeholder responses:

1. What question(s) about the diagnosis of pituitary adenomas requiring surgery would you like to see answered by research?
2. What question(s) about the surgical treatment of pituitary adenoma would you like to see answered by research?
3. What question(s) about the long-term care and follow-up after pituitary adenoma surgery would you like to see answered by research?
4. What other question(s) about pituitary adenoma surgery that do not fit into the above categories would you like to see answered by research?

Research uncertainties will be grouped thematically, and duplicates will be removed. Out-of-scope responses will be reviewed by the steering group and will be excluded from further analysis if all members agree. Analysis will be in the form of inductive thematic analysis where themes will be generated directly from the data by identifying recurring topics and ideas. The themes will then be created into distinct candidate research uncertainties^16,17^. A review of the literature will be performed to identify which of these candidate research uncertainties are unanswered. These unanswered research uncertainties will be compiled and organised into parent categories (diagnosis, management, follow-up, other).

### Round 2 – online survey

In scope unanswered research uncertainties from survey round 1 will be presented in the round 2 survey. Participants will be asked to select their top 10 research uncertainties. A link to the online survey will be disseminated through organisations described previously and directly to those who had provided contact details in the round 1 survey. Round 2 will be open to participation from individuals who did not participate in round 1. Research uncertainties will be ranked based on the number of times they were included in participants’ list of top 10 research uncertainties. Ranked lists will also be generated for stakeholder group. The top 20-30 research uncertainties will be brought forward to round 3.

### Round 3 - online consensus meeting

The aim of this meeting will be to establish the top 10 research uncertainties from the established shortlist of questions via a face-to-face consensus meeting. The final prioritisation workshop will be attended by 10 representatives. Representatives will be carefully selected to ensure equal representation across all stakeholder groups in the workshop in a 1:1 ratio of healthcare professionals to service users. Before the workshop, each participant will be asked to review the shortlist of research uncertainties and prioritise them. Each question will have interim priority setting data on the back, including how many survey respondents from each stakeholder group ranked the question in their top ten.

The workshop will consist of one round of discussion. An information specialist will be assigned to the working group and will ask the group to rank their priorities and rationale for their choice. This will enable each stakeholder to voice their reasons for or against each priority. The final top 10 priorities will then be established.

#### COS study design

This study and protocol have been designed and written in accordance with the Core Outcome Measures for Effectiveness Trials (COMET) Handbook, Core Outcome Set-STAndards for Development (COS-STAD) and Core Outcome Set-STAndardised Protocol (COS-STAP) statements^18,19^. Results will be reported according to the Core Outcome Set Standards for Reporting (COS-STAR)^11^guidelines. A multi-round online Delphi survey will be conducted on the online survey tool-*Qualtrics*. A study timeline can be found in Figure 2.

**Figure 2:**
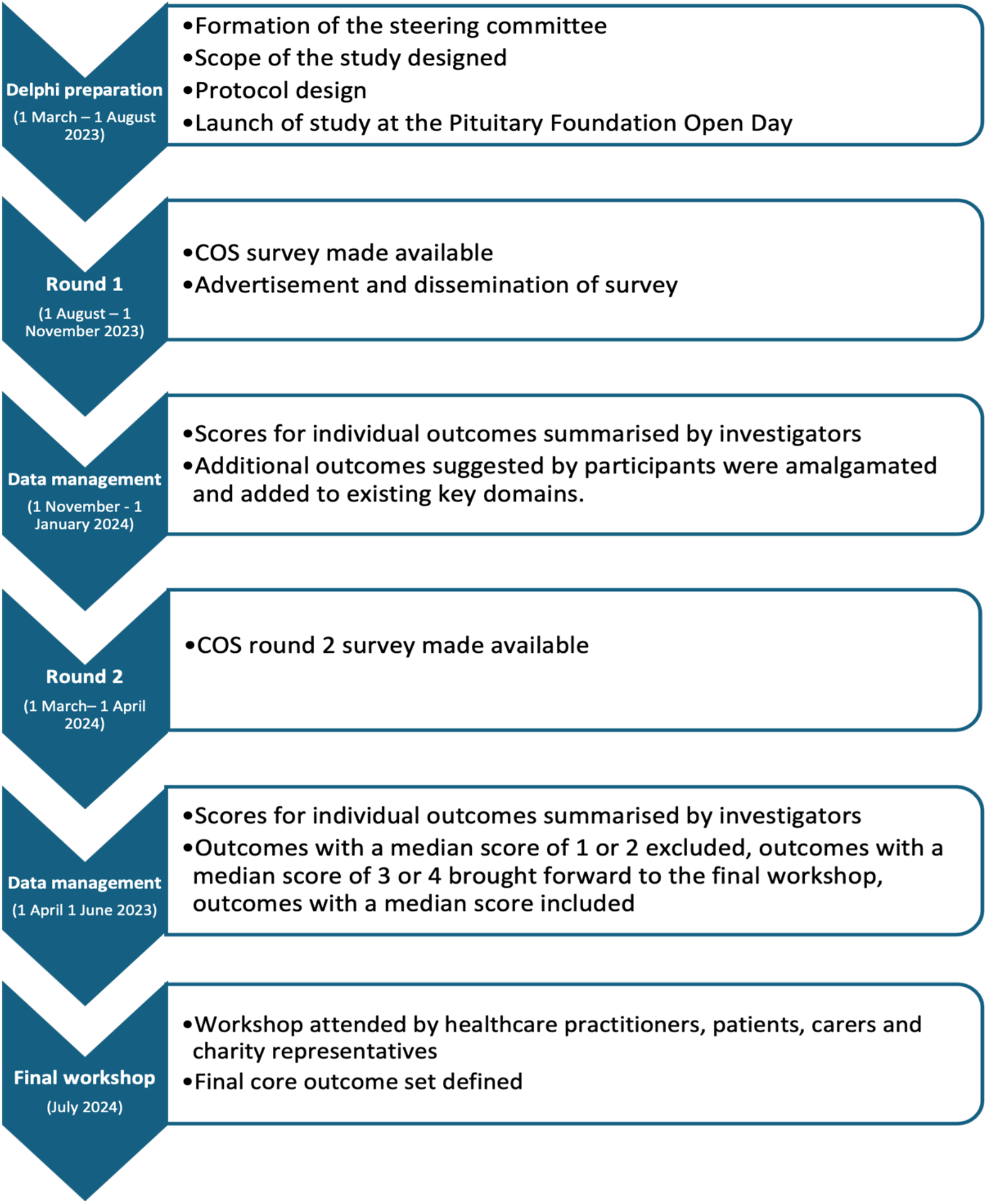
COS study timeline

### Round 1- online survey

The initial list of outcomes for this study were identified from (i) a systematic review which reported outcomes after transsphenoidal surgery for pituitary adenoma, and (ii) a study which developed and validated a patient reported outcome measure (PROM) for patients with pituitary adenoma undergoing transsphenoidal surgery^20,21^.

In the first-round, participants will be presented with a list of outcomes grouped into key domains^20,21^. Participants will then be asked to rate each outcome according to how strongly they agree or disagree it should be included in the core outcome set using a 5-point scale. On the 5-point scale 1 is ‘strongly disagree’, 2 is ‘disagree’, 3 is ‘neutral’, 4 is ‘agree’ and 5 is ‘strongly agree’. Participants will be able to provide a rationale for their rating and suggest additional outcomes for consideration. The consensus definition used throughout this study is outlined in Table 1.

**Table 1:** Consensus definition.

|  | Rounds 1 and 2 |  | Consensus meeting |  |
| --- | --- | --- | --- | --- |
| Category | Definition | Action | Definition | Action |
| Consensus to include | Median score of 5 (strongly agree). | After round 1: outcome included in next Delphi round. After round 2: outcome provisionally included in the final COS. | As per round 1 and 2 | Outcome included in final COS. |
| Consensus to exclude | Median score of 1– (strongly disagree) or 2 (disagree) | After round 1: outcome include in the next Delphi round. After round 2: outcomes provisionally excluded from the final COS. | As per round 1 and 2 | Outcome not included in final COS. |
| No consensus | Median score of 3 (neutral) or 4 (agree) | After round 1: outcome included in next Delphi round. After round 2: outcome brought to the round 3 consensus meeting for discussion. | As per round 1 and 2 | Round of discussion for consensus to be reached. |

Data from the first round will be analysed by calculating the overall median and interquartile range (IQR) for each outcome. Additional outcomes which were suggested by participants will be amalgamated and added to existing key domains. Additional outcomes will be excluded if considered not within scope by the management committee.

### Round 2 – online survey

Participation in round two will be restricted to those who completed round one. Participants will again be presented with a list of each outcome, though this time each outcome will be accompanied by the summarised group score (median and IQR) from round 1. Participants will be asked to re-score each outcome on the same 5-point scale while considering the summarised group scores from round one.

Data from round two will be analysed by calculating the overall median and IQR for each outcome. Outcomes with a median score of 1 (strongly disagree) or 2 (disagree) will be provisionally excluded from the final core outcome set. Outcomes with a median score of 5 (strongly agree) will be provisionally included in the final core outcome set. Outcomes with median scores 3 (neutral) or 4 (agree) will be brought forward for discussion in round 3.

### Round 3 – online consensus meeting

The final round will involve a live consensus meeting to discuss borderline outcomes (median score 3 or 4) after round two scoring. The discussion will determine which of the borderline outcomes should be included in the core outcome set. Outcomes for inclusion after round two (median score 5) will also be ratified in the live meeting. Outcomes excluded after round two (median scores 1 or 2) will also be discussed to allow an opportunity for stakeholders to express any concern regarding their exclusion.

## Data Availability

All data produced in the present study are available upon reasonable request to the authors

## Dissemination

Study results will be disseminated through international conference presentation, publication in an open access peer-reviewed journals, relevant societies and charities, and via social media.

## Acknowledgements

We would like to thank the National Brain Appeal, Wellcome / EPSRC Centre for Interventional and Surgical Sciences (WEISS), University College London Hospitals, University College London, The University of Cambridge and the Pituitary Society for supporting the research.

